# Genetic liability for schizophrenia and childhood psychopathology in the general population

**DOI:** 10.1101/2020.05.01.20086215

**Authors:** Laurie J. Hannigan, Ragna Bugge Askeland, Helga Ask, Martin Tesli, Elizabeth Corfield, Ziada Ayorech, Øyvind Helgeland, Per Magnus, Pål Rasmus Njølstad, Anne-Siri Øyen, Camilla Stoltenberg, Ole A. Andreassen, George Davey Smith, Ted Reichborn-Kjennerud, Alexandra Havdahl

## Abstract

**Background:** Genetic liability for schizophrenia is associated with psychopathology in early life. It is not clear if these associations are time-dependent during childhood, nor if they are specific across different forms of psychopathology.

**Methods:** Using genotype and questionnaire data on children (N = 15,105) from the Norwegian Mother, Father, and Child Cohort Study (MoBa), we tested associations between schizophrenia polygenic risk scores and measures of childhood emotional and behavioural problems for developmental stability and domain specificity. We then sought to identify symptom profiles – across development and domains – associated with elevated schizophrenia polygenic liability.

**Outcomes:** We found evidence for developmental stability in associations between schizophrenia polygenic risk scores and emotional and behavioural problems, with the latter being mediated via the rate of change in symptoms between 18 months and 5 years specifically (β_slope_ = 0.032; 95% CI 0.007 – 0.057). At age 8, associations with emotional and behavioural psychopathology were found to be better explained by a model of symptom-specific polygenic risk score effects, rather than effects mediated via a general “p” factor or by domain-specific factors. Overall, individuals with higher schizophrenia polygenic risk scores were more likely (OR= 1.310 [95% CIs: 1.122-1.528]) to have increasing behavioural and emotional symptoms in early childhood, followed by relatively elevated symptoms of conduct disorder, oppositional defiant disorder, hyperactivity and inattention in middle childhood.

**Interpretation:** Schizophrenia-associated alleles are linked to specific patterns of early-life psychopathology. The associations are small, but findings of this nature can help us better understand the developmental emergence of schizophrenia.

**Funding:** Laurie Hannigan, Ziada Ayorech, and Alexandra Havdahl were supported by grants from the South-Eastern Norway Regional Health Authority (2018059, 2019097 and 2018058, respectively). Ragna Bugge Askeland and Elizabeth Corfield were supported by a grant from the Norwegian Research Council (274611). Ted Reichborn-Kjennerud was supported by a grant from the Research Council of Norway (274611). Pål Rasmus Njølstad was funded by the ERC AdG SELECTionPREDISPOSED (#293574), the Stiftelsen Kristian Gerhard Jebsen, the Trond Mohn Foundation, the Norwegian Research Council (#240413/F20), the Novo Nordisk Foundation (#54741), the University of Bergen, and the Western Norway health Authorities (Helse Vest; PERSON-MED-DIA and #911745). Ole A. Andreassen was supported the Research Council of Norway (229129; 213837; 248778; 223273; 249711); the South-East Norway Regional Health Authority (2017-112); KG Jebsen Stiftelsen (SKGJ) and H2020 grant # 847776 CoMorMent. George Davey Smith works in the Medical Research Council Integrative Epidemiology Unit at the University of Bristol, which is supported by the Medical Research Council (MC_UU_00011/1). This work was partly supported by the Research Council of Norway through its Centre of Excellence funding scheme, project number 262700.

**Research in context:** *Evidence before this study:* Genetic liability to schizophrenia, conferred incrementally by many genetic variants of small effect, is associated with various forms of psychopathology – both symptoms and diagnoses – in the general population during adulthood. To get an overview of the relevant evidence for how genetic liability to schizophrenia manifests in psychopathology in *childhood*, we performed a systematic search of the published literature using the Ovid MEDLINE and PsychINFO databases, for English-language peer-reviewed journal articles published prior to 28 January 2020. We found 7 studies of core relevance (i.e., containing assessment of symptoms or diagnoses of psychopathology in pre-adolescent children), with a further 9 studies examining outcomes potentially related to psychopathology (such as brain structure, cognitive performance, and social functioning). Of the 7 core studies, 3 used clinical samples to demonstrate that polygenic risk scores for schizophrenia are higher among children with ADHD diagnoses than controls, and among cases of the rare child-onset form of schizophrenia than their healthy siblings. The remainder of studies all examined symptoms of psychopathology among children in the general population, finding modest but robust associations of schizophrenia polygenic risk scores with emotional and behavioural problems measured from 3 years of age, as well as with symptoms of depression, ADHD, anxiety, oppositional defiant disorder and conduct disorder in middle childhood.

*Added value of this study:* In this study, we present a set of analyses designed to improve our understanding of the nature of associations between schizophrenia risk alleles and childhood psychopathology. Specifically, we employ an approach that aims not just to quantify, but also to explore how the effects of schizophrenia risk manifest across childhood, and across different domains of psychopathology. We find evidence that effects of schizophrenia polygenic risk scores on symptoms of emotional and behavioural problems in early childhood are stable, influencing the overall level and rates of change in symptoms, rather than age-specific (i.e., transient or developmental). We also find evidence of specificity in the effects of schizophrenia polygenic risk scores on different domains of psychopathology in 8-year-old children. Overall, we find that higher schizophrenia polygenic risk scores are associated with a developmental symptom profile comprising elevated and increasing symptoms of behavioural problems and increasing levels of emotional problems in early childhood, as well as particularly elevated symptoms of conduct disorder, inattention, hyperactivity, and oppositional defiant disorder in middle childhood.

*Implications of all the available evidence:* Findings of our study align with a growing body of evidence that the effects of schizophrenia risk alleles on psychopathology begin early in life, and influence the likelihood of children experiencing difficulties across development. While previous work has largely found similar effects of schizophrenia polygenic risk scores across different domains of childhood psychopathology, indicating that such effects may be mediated by a hypothetical latent ‘general psychopathology’ or ‘p’ factor, our results suggest that domain- and even symptom-level specificity may emerge by middle childhood. We may be able to improve our understanding of processes underpinning the emergence of schizophrenia later in life by paying attention to nuances in the ways that genetic risk for schizophrenia manifests across childhood and into adolescence.

## Introduction

Genetic risk for schizophrenia is highly polygenic (1,2), representing an additive combination of many common genetic variants with small effects. Large-scale genome-wide association studies (GWAS) (3) discover these effects, with currently more than 176 genetic loci identified as conferring risk for schizophrenia (4). Effect sizes and standard errors from GWAS can be combined across many variants in a polygenic risk score (PRS) (5), to examine how common genetic liability for schizophrenia is associated with traits and behaviours in the general population (6).

Schizophrenia is most commonly diagnosed in late adolescence or early adulthood (7). PRS-based approaches can be used to study early-life manifestations of genetic risk for schizophrenia, with the potential to inform as to how and why the disorder emerges later in life. Previous studies have shown that genetic liability for schizophrenia is modestly associated with a range of childhood outcomes, including infant neuromotor development (8), early neurocognitive and behavioural development (9), sleep problems (10) and social cognition (11) – as well as with measures of psychopathology (including symptoms of anxiety, depression, attention deficit hyperactivity disorder, and conduct problems) across childhood (12–15). These associations appear to persist into adolescence (16,17) as well as potentially diversifying further (e.g., into disordered eating (18) and cannabis use (19)).

Although it seems clear that early life manifestations of schizophrenia genetic liability across the psychopathological “phenome” are relatively diverse, it is not well understood how this diversity arises. For example, it could be that schizophrenia-associated genes have transient, developmentally-varying effects; or that early effects in specific domains trigger developmental cascades that encompass a wider range of behaviours. It could simply be the case that these genes have highly generalized effects on behaviour; or that the environmental and maturational restrictions of childhood make their expression more diffuse than is observed in adulthood. To narrow down the various possible explanations, it is necessary to investigate associations in two additional dimensions: developmental time, and phenotypic space. That is, to ask to what degree effects are developmentally stable (influencing behaviour consistently across development) *vs*. age-specific (transient or emerging at a specific point), and to explore how broadly *vs*. selectively they influence different behaviours and symptoms.

In the current study, we use data from the population-based pregnancy cohort: the Norwegian Mother, Father, and Child Cohort Study (MoBa) (20) to investigate the extent to which manifestations of genetic liability for schizophrenia in childhood emotional and behavioural problems are: i) developmentally stable *vs*. unstable (age-specific); and ii) domain-general *vs*. domain-specific. Additionally, we investigate genomic prediction, using schizophrenia PRS, of individuals’ latent symptom profiles across development and a broad range of symptom domains.

The analyses we present are exploratory, with the aim of identifying the most parsimonious of a set of models describing the covariation between schizophrenia PRS and questionnaire measures of childhood emotional and behavioural problems. However, we also had two broad hypotheses. First, we expected developmentally-stable associations between schizophrenia-associated genetic variants and symptoms of psychopathology across early and middle-childhood, based on both work with schizophrenia PRS (12,13,15) and findings of the stable genetic influence on childhood psychopathology (21). Second, we expected associations later in childhood to be primarily general, mediated via a latent, general *“p*” (for *“psychopathology’)* factor (22). The *“p*”factor model offers a parsimonious explanation for comorbidity and shared genetic influence between symptom domains (23,24) and schizophrenia PRS have been shown to explain variance in the latent *“p”* component of the model in childhood (25) and adolescence (26).

## Methods

### Study sample

The Norwegian Mother, Father and Child Cohort Study (MoBa) is a population-based pregnancy cohort study conducted by the Norwegian Institute of Public Health (27). Participants were recruited from all hospitals and obstetric units in Norway during 1999-2008. The women consented to participation in 41% of the pregnancies. The cohort now includes 114,500 children, 95,200 mothers and 75,200 fathers. The current study is based on version 12 of the quality-assured data files released in January 2019.

The establishment and data collection in MoBa were previously based on a license from the Norwegian Data Protection Agency and approval from The Regional Committee for Medical Research Ethics (REK) and is now based on regulations under the Norwegian Health Registry Act. The current study was approved by REK (2016/1702).

Blood samples were obtained from children (umbilical cord) at birth. Genotyping of the entire MoBa cohort is ongoing. We used genotype data from children in 17,000 randomly selected trios, and after quality control the analytic sample was a genotyped sub-set (N = 15,105) of children (20). Details about the processing of the genetic data are outlined in the Appendix.

### Measures

We used measurements of children’s symptoms of emotional and behavioural psychopathology from maternal questionnaires collected when children were aged 18 months, 3 years, 5 years, and 8 years. Fifteen items from the Child Behaviour Checklist (CBCL) (28), which could be sub-divided into emotional problems (5 items) and behavioural problems (10 items), were included for longitudinal analyses across early childhood (18 months to 5 years). Three instruments measuring six domains of emotional and behavioural psychopathology in middle childhood (at the 8-year data collection) were included for the specificity analyses. The 13-item Short Mood and Feelings Questionnaire (sMFQ) (29) measured symptoms of depression, a 5-item short form of the Screen for Child Anxiety Related Disorders (SCARED) (30) measured symptoms of anxiety, and a 34-item version of the Rating Scale for Disruptive Behaviour Disorders (RS-DBD (31)) measured symptoms of conduct problems (CD), oppositional defiant disorder (ODD), hyperactivity, and inattention. Details about measure selection and psychometrics are in the appendix.

### Polygenic risk scores

We calculated PRS for schizophrenia using PRSice2 (32), based on European samples from the most recent (2) Psychiatric Genomics Consortium schizophrenia genome-wide association study (GWAS). PRS can be calculated using effect estimates for all variants in common between the discovery (i.e., GWAS) sample and target sample, for variants whose effects in the GWAS had a p-value below a specified threshold. Typically, PRS are created at a range of p-value thresholds (0-1), reflecting the expectation that the polygenic signal will gradually increase as more variants (either with weaker effects or a lower frequency in the population) are included, up until the point additional variants contribute only statistical noise to the score. We opted to use the thresholds from the polygenic scoring analysis presented in the original GWAS (2). These were: *p* < 0.001, *p* < 0.01, *p* < 0.05, *p* < 0.1, *p* < 0.2, *p* < 0.5, *p* < 1. Full details of the parameters and procedure used in generating the PRS are in the Appendix.

### Analyses

#### Developmental modelling

Latent growth models were used to explore variability in the development of emotional and behavioural problems between the ages of 18 months and 5 years in the context of genetic liability for schizophrenia. In these models, variance in observed variables is explained by two latent growth factors: a latent intercept, which is specified to load equally at all waves (here at 1.5, 3, and 5 years), and a latent slope, loadings for which are fixed proportional to their temporal distance from the first wave of measurement (here the loadings are 0, 1.5, and 3). For each schizophrenia PRS (based on the different thresholds), we formally compared the fit of models specifying an effect on growth parameters (intercept and slope) *vs* age-specific residuals in emotional and behavioural problems. This model fitting indicated to what extent the association between schizophrenia PRS and emotional/behavioural problems is developmentally stable (i.e., via latent growth factors), age-specific (i.e., via residuals), or null. Informal comparisons were also made to ascertain whether stable effects could be primarily ascribed to either the latent intercept or slope factor (for full details of model fitting and comparison strategy, see appendix).

#### *“p”* factor modelling

Next, we specified models describing covariation among items from the 8-year psychopathology variables with 6 domain-specific factors and one overall “p” factor. We compared their fit to the data when schizophrenia PRS were allowed to influence, respectively: symptom-specific residuals, domain-specific factors, the domain-general “p” factor, as well specified to have no effect. All versions of a given model were nested, so formal tests of fit (χ^2^ difference tests) were used to ascertain the best-fitting model. For both the developmental and “p” factor modelling, we examined the average R^2^ value among the outcomes and present results for the PRS threshold at which R^2^ was maximised.

#### Latent profile analysis

Finally, we incorporated both the developmental modelling and *“p”* factor modelling approaches into a latent profile analysis. In latent profile analysis, a categorical latent variable is used to assign individuals in a sample into one of a pre-specified number of profiles based on their pattern of scores across multiple observed continuous variables. For our analysis, profile membership was informed by individuals’ estimated scores on the continuous latent growth factors from the emotional and behavioural problems latent growth models, and their observed scores on compiled scale versions of the 6 psychopathology domains measured at 8 years. We then included schizophrenia PRS as predictors of profile assignment. The full model is displayed in Figure 1. We specified models with 2, 3, 4, 5, and 6 profiles respectively, and assessed them using a combination of standard criteria (Vuong–Lo–Mendell–Rubin likelihood ratio test (33,34), entropy, fit indices).

**Figure 1.**
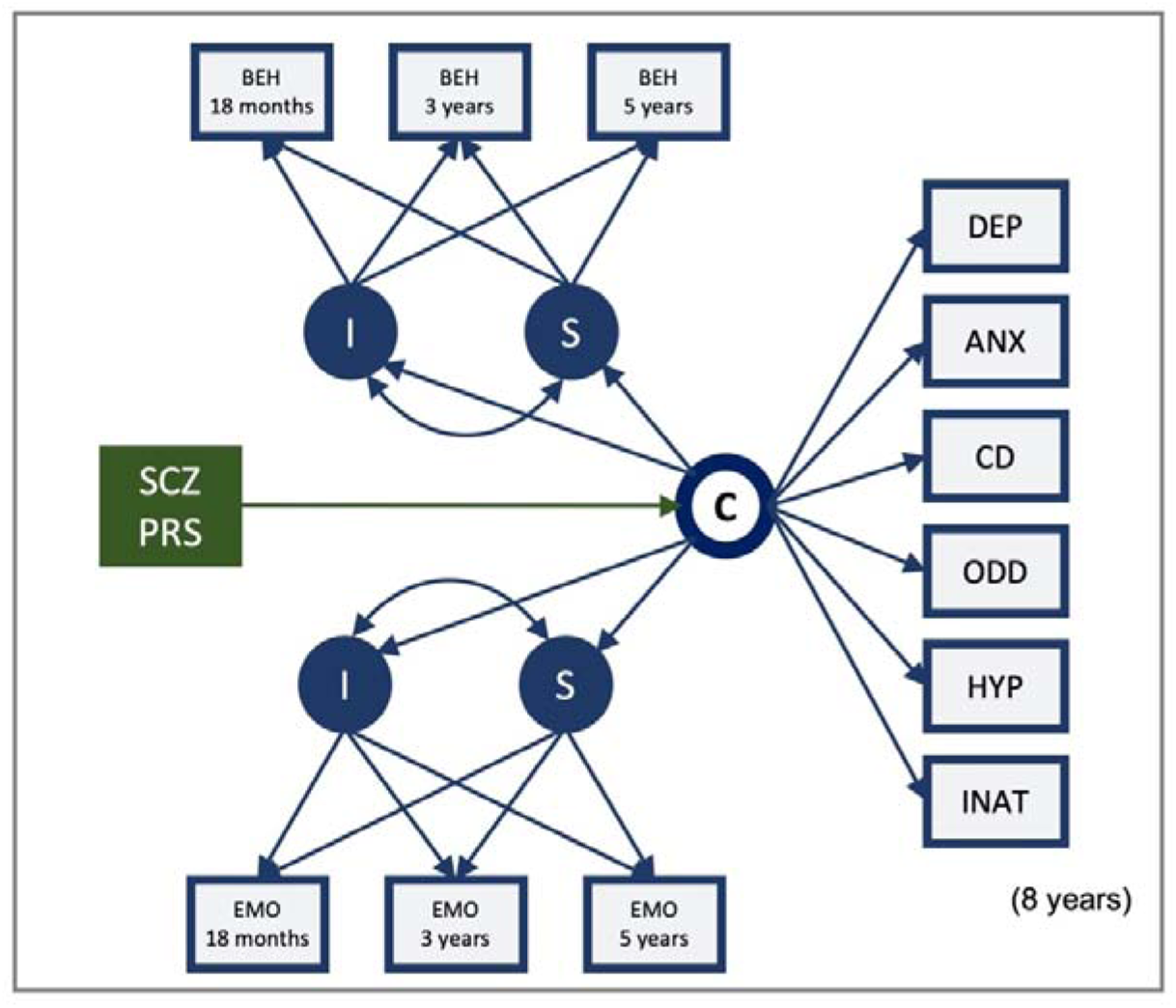
Latent profile analysis to ascertain developmental and domain-specific profiles associated of psychopathology symptoms associated with schizophrenia genetic effects.

*Note-boxes represent observed variables and circles model-estimated latent variables; I = intercept factor, which loads equally on observed variables at all waves; C = categorical latent variable, sub-dividing the sample into a specified number of classes (we tested models with 2, 3, 4, 5, and 6) according to values on: S = slope factor with loadings 0, 1.5, 3.5, corresponding to temporal distance from the first wave of measurement; EMO = CBCL emotional problems symptoms; BEH = CBCL behavioural problems symptoms; DEP = sMFQ depressive symptoms; ANX = SCARED anxiety symptoms; CD = RS-DBD conduct disorder symptoms; ODD = RS-DBD oppositional defiant disorder symptoms; HYP = RS-DBD hyperactivity (ADHD) symptoms; ODD = RS-DBD inattention (ADHD) symptoms; 8-year observed variables and internalizing/externalizing intercept/slope variables, respectively, are inter-correlated within class (paths omitted from diagram for clarity)*

All modelling for the first two parts of the analyses was carried out in R version 3.4.4 using the *lavaan* package (35) version 0.6.3, and the latent profile analysis required the *Mplus* software (36) version 8.1 via the R package *MplusAutomation* (37) version 0.7.3, as *lavaan* does not support categorical latent variables at present. All models included sex as a covariate. Further details are available in the appendix, and code for the modelling is openly available at https://github.com/psychgen/scz-prs-psychopathol-dev.

## Results

Descriptive statistics for all study variables are available in **Table 1**, alongside an exploration of the extent of selection bias among our genotyped sub-sample of MoBa and of selective attrition with respect to the longitudinal analyses (both of which were observed at low levels) is included in the Appendix.

**Table 1.** Descriptive statistics for main study variables (scale-level only)

| Measure | Age (yrs) | N | Mean | SD | Min | Max |
| --- | --- | --- | --- | --- | --- | --- |
| Behavioural problems (CBCL) | 1.5 | 11552 | 3.87 | 2.21 | 0 | 16 |
|  | 3 | 9369 | 3.78 | 2.38 | 0 | 16 |
|  | 5 | 7274 | 2.42 | 2.23 | 0 | 15 |
| Emotional problems (CBCL) | 1.5 | 11549 | 1.26 | 1.18 | 0 | 8 |
|  | 3 | 9370 | 1.33 | 1.35 | 0 | 9 |
|  | 5 | 7272 | 1.03 | 1.26 | 0 | 10 |
| Depressive symptoms (sMFQ) | 8 | 7298 | 1.78 | 2.36 | 0 | 23 |
| Anxiety symptoms (SCARED) | 8 | 7310 | 0.75 | 1.49 | 0 | 16 |
| Conduct disorder symptoms (RS-DBD) | 8 | 7303 | 3.46 | 3.86 | 0 | 27 |
| Oppositional defiant disorder symptoms (RS-DBD) | 8 | 7304 | 4.82 | 4.00 | 0 | 27 |
| Hyperactivity symptoms (RS-DBD) | 8 | 7300 | 3.40 | 3.10 | 0 | 22 |
| Inattention symptoms (RS-DBD) | 8 | 7307 | 1.01 | 1.20 | 0 | 10 |

*Note: CBCL = Child Behavior Checklist; sMFQ = Short Mood and Feelings Questionnaire; SCARED = Screen for Child Anxiety Related Disorders; RS-DBD = Rating Scale for Disruptive Behaviour Disorders*

### Schizophrenia genetic liability and developmental stability in childhood psychopathology

Linear growth models provided an acceptable fit to the data from the measures of emotional and behavioural problems (for details, see appendix). We found evidence of associations between schizophrenia genetic liability and emotional and behavioural problems, with the null model rejected at most p-value thresholds in each domain. Moreover, we observed evidence in favour of developmentally stable rather than age-specific associations, with models incorporating PRS effects on the latent growth factors preferred in both domains (see appendix for genetic model-fitting details). For emotional problems, we could not formally distinguish whether these effects were primarily mediated via the intercept (i.e., predicting the overall level of symptoms) or slope (i.e., rate of change in symptoms over time) in the best-fitting model, and point estimates were similar for both (β_intercept_ = 0.019, 95% CI: −0.005-0.043; β_slope_ = 0.019, 95% CI: −0.010-0.048; PRS threshold: *p*<0.1). In contrast, for behavioural problems, these effects were mediated via the slope factor alone at the most predictive threshold (β_slope_ = 0.032; 95% CI 0.007-0.057; threshold: p<1), meaning that schizophrenia PRS predicted rate of change in symptoms of behavioural problems across early childhood. However, in both cases the associations were small, explaining ~0.1% variance in the growth factors. Parameter estimates and 95% CIs for best-fitting models are displayed in the Appendix.

### Schizophrenia genetic liability and phenotypic specificity in childhood psychopathology

A structural model of domain-specific factors and one overarching “p” factor provided a good fit to item-level data from the 8-year psychopathology measures (CFI=0.98, TLI=0.97, RMSEA=0.04). Details of the model fitting process are presented in the appendix. We found that schizophrenia PRS were significant predictors of variability in symptoms of psychopathology at 8 years, and that this prediction was maximised at the PRS threshold including all variants (*p*<1). At this threshold, neither the model incorporating a PRS effect on the general “*p*” factor nor the model incorporating domain-specific effects fit the data sufficiently well to be accepted. Instead, the preferred model allowed the PRS to influence item-specific residuals – i.e., explaining symptom-specific variation. **Figure 2** shows standardized beta coefficients for these symptom-level relationships.

**Figure 2.**
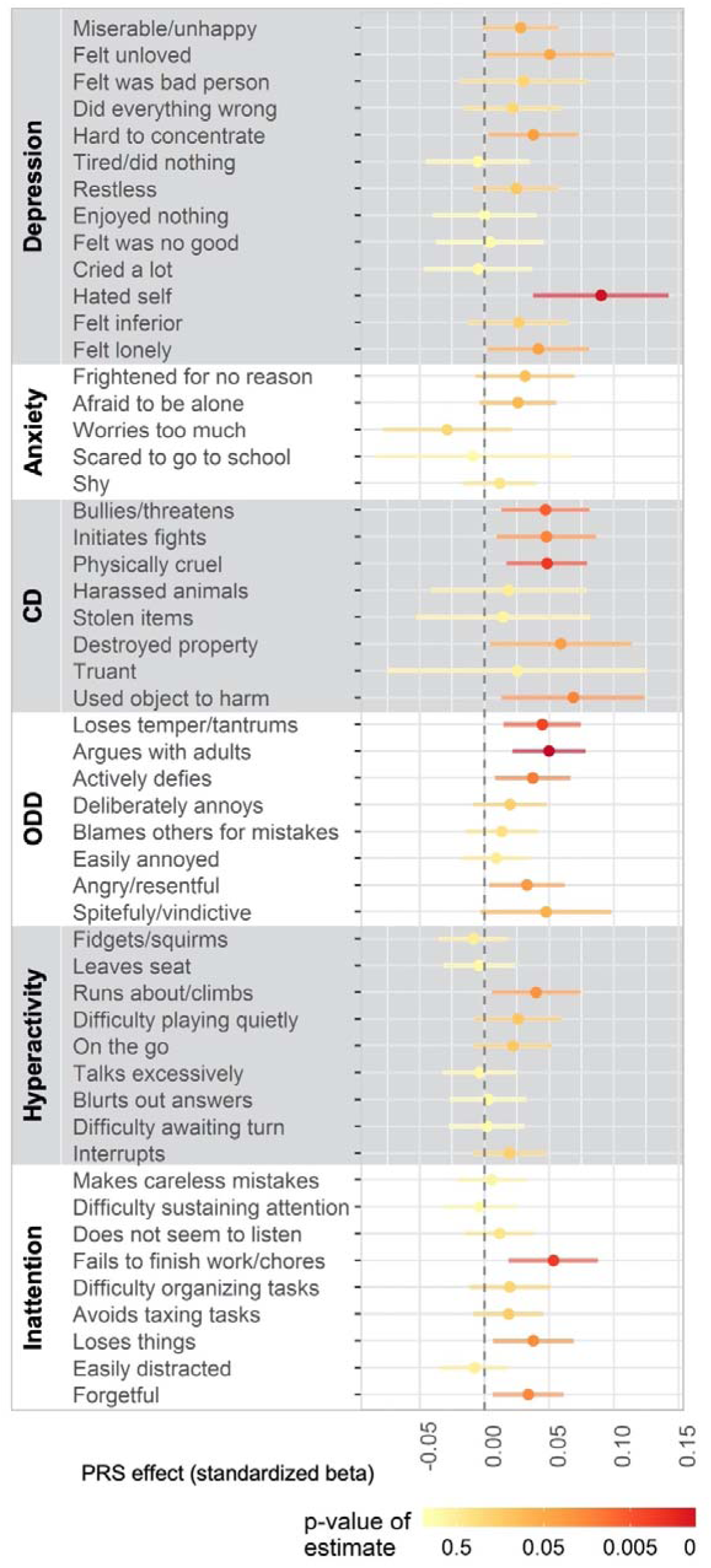
Item-level heterogeneity of schizophrenia polygenic risk score associations with 8-year childhood psychopathology symptoms.

*Note-Bars indicate 95% confidence intervals; PRS threshold p<1 (i.e., all variants)*

The pattern of results in **Figure 2** show that there is substantial heterogeneity, in terms of schizophrenia PRS associations, between symptoms – even within domains. This is particularly evident for symptoms within the depression and inattention domains. As with the developmental modelling, effect sizes were very small, with PRS explaining a maximum of 0.4% variance in symptoms. Parameter estimates and 95% CIs for the alternate versions of the model, which were preferred for PRS comprising variants significant only at less stringent p-value thresholds, are presented in the appendix.

### Schizophrenia genetic liability predicting developmental symptom profiles

Scores from all scales (CBCL measures of emotional and behavioural problems at 18 months, 3 years, and 5 years and six 8-year psychopathology domains) were incorporated into a single model for the latent profile analysis. Fit statistics for these models are presented in the appendix. Entropy (reflecting the overall certainty with which individuals could be assigned to symptom profiles) was good (~0.8) for each version of the model, but the Vuong–Lo–Mendell–Rubin test statistic for the 5-profile model indicated that it offered no significant improvement of fit on the simpler 4-profile model.

**Figure 3** shows the 4 symptom profiles. Individuals assigned to profile 1 (5.7% of sample) had increasing symptoms of emotional problems between 18 months and 5 years and elevated anxiety symptoms at age 8. Profile 2 (7.9%) was characterised by moderate, stable behavioural problems symptomatology in early childhood, and moderate symptoms of ODD, hyperactivity, and inattention at 8 years. Profile 3 was considered the normative symptom profile, as a large majority (84.7%) could be assigned to it, and it was characterised by decreasing problems developmentally and low levels of symptoms at 8 years. Profile 4 was the least populous (1.8% of the sample) and most symptomatic profile, characterised by increasing/elevated symptoms of behavioural problems and increasing symptoms of emotional problems preceding relatively elevated symptoms of conduct disorder, ODD, hyperactivity and inattention at 8 years.

**Figure 3.**
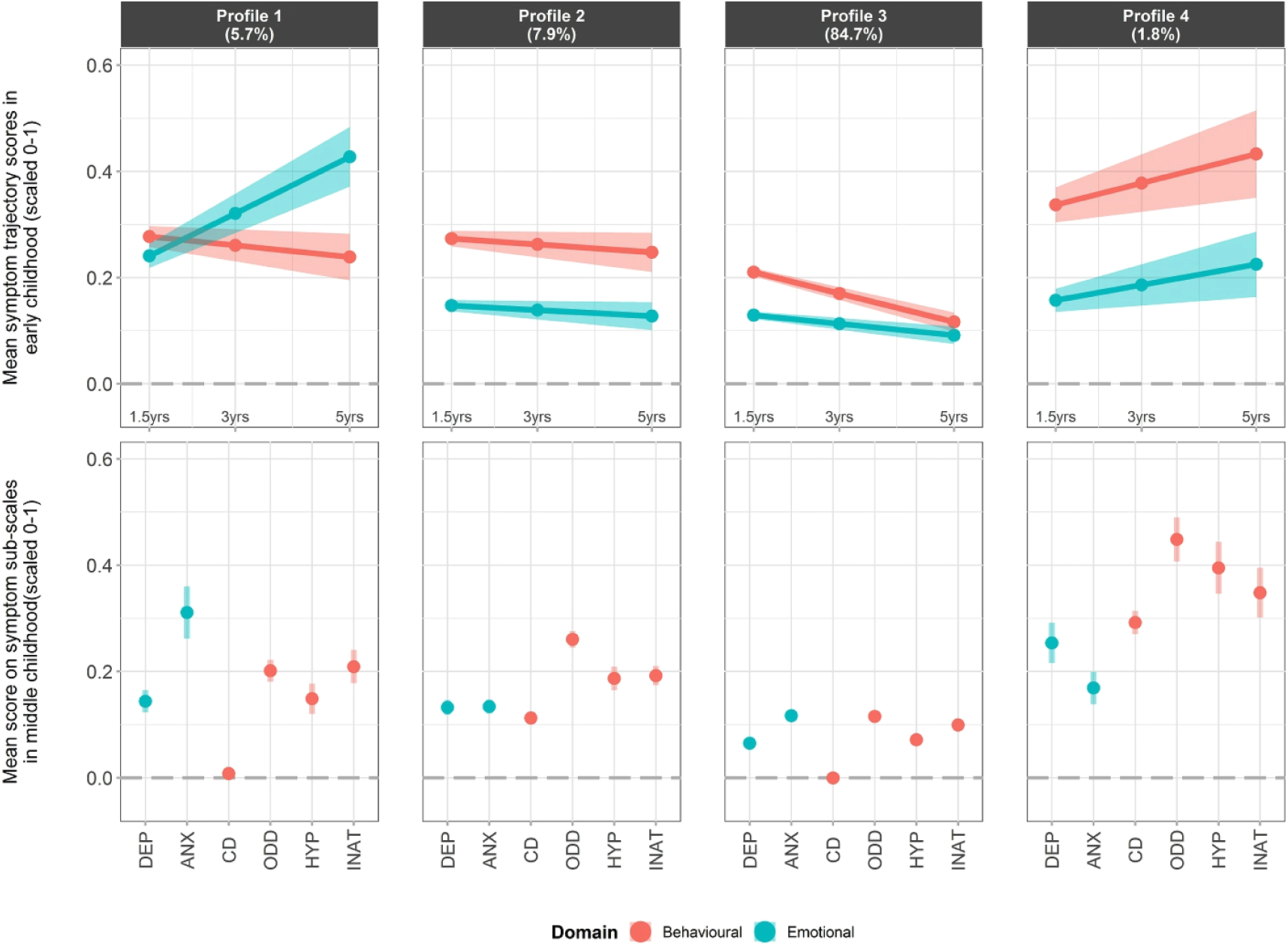
Symptom profiles from latent profile analysis of symptoms of psychopathology across development and domain.

*Note-Bars/bands indicate 95% confidence intervals; %s in header indicate the proportion of the sample best classified in each profile; DEP = sMFQ depressive symptoms; ANX = SCA, anxiety symptoms; CD = RS-DBD conduct disorder symptoms; ODD = RS-DBD oppositional defiant disorder symptoms; HYP = RS-DBD hyperactivity (ADHD) symptoms; ODD = RS-D inattention (ADHD) symptoms; 8-year observed variables and internalizing/externalizing intercept/slope variables, respectively, are inter-correlated within class (paths omitted from diag, for clarity)*

The probabilities of classification into profiles 1, 2, and 4 relative to the normative profile are shown in **Figure 4** as a function of PRS (at the *p* <0.05 threshold), alongside odds ratios of PRS on probability of classification into a given profile rather than the normative profile. Profile 4 was better defined than profiles 1 or 2 (the density of high probabilities of assignment into this profile in the bottom-left panel of **Figure 4** is greater than in the two panels above, and there is greater separation between individuals classified in this profile and those with normative profile). The average PRS was higher in profile 4 than in all other profiles, and indeed individuals with higher PRS were more likely to be assigned to profile 4 than the normative profile (OR= 1.310 [95% CIs: 1.122-1.528], bottom-right of figure). There was no strong evidence of associations between schizophrenia PRS and probability of assignment to either of the other profiles (relative to the normative profile).

**Figure 4.**
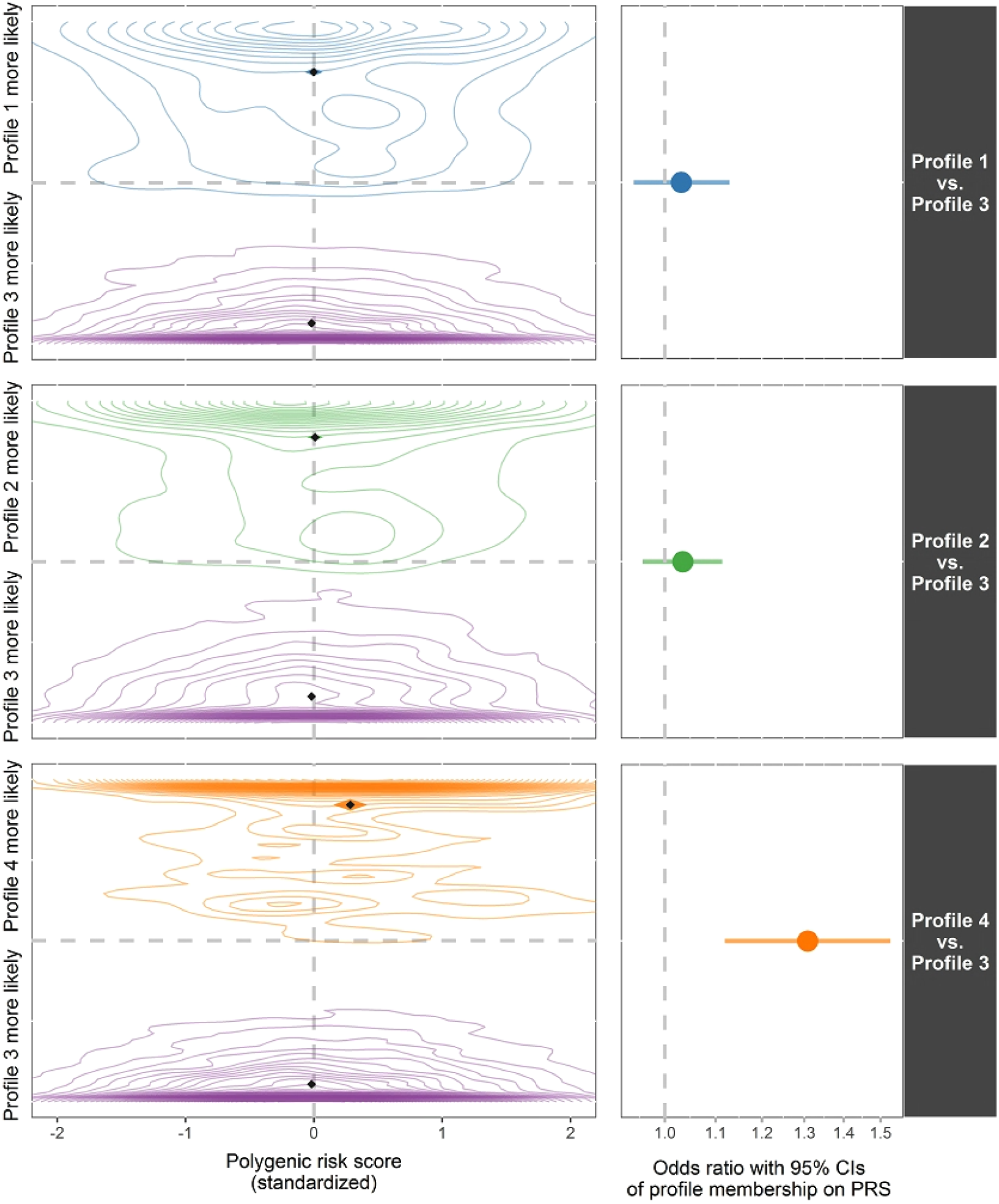
Density of relative probabilities of classification into the different profiles by schizophrenia PRS and odds ratios for prediction of symptom profile classification by PRS.

*Note-contour plots show probabilities of classification into a specific profile (1, 2, or 4) relative to probabilities of classification into the normative profile (3) for individuals ultimately assigned to each of these profiles (indicated by colour coding), as a function of PRS; diamond marker indicates within-profile means (both classification probability and PRS) and the shaded region around them shows the 95% CIs in either dimension (i.e., vertically for probability, horizontally for PRS); profile 3 is used as reference category for logistic regression; PRS threshold used is p<0.05, selected as it maximizes the OR for profile 4 vs*. *profile 3; but the pattern of results is consistent at other thresholds (see Appendix)*

## Discussion

The results indicate that associations between schizophrenia risk-associated alleles and symptoms of early childhood (18 months to 5 years) psychopathology tend to be developmentally stable, rather than transient or emergent. In middle childhood (8 years) we found that, contrary to expectations, associations between schizophrenia genetic risk and symptoms of psychopathology may be better explained by symptom-specific effects than by prediction of a latent *“p”* factor indexing general psychopathology. Finally, we showed that by combining information about symptoms of psychopathology across time and domains, associations with schizophrenia genetic liability can be summarised in the form of a characteristic symptom profile.

Our results are consistent with findings of widespread associations between schizophrenia genetic liability and symptoms of childhood psychopathology. There are a number of possible explanations for such associations. They may arise because of *pleiotropy;* that is, direct effects of schizophrenia-related genetic variants on other behaviours and symptoms. They may represent early manifestations of schizophrenia liability, where environmental or maturational restrictions mean that the classic “schizophrenia phenotype” (and its sub-clinical analogues) cannot be fully expressed until adolescence or later. A further alternative is that (some) childhood manifestations of genetic liability for schizophrenia are actually part of a causal process in the development of the disorder. This could involve intrinsic, developmental processes (e.g., specific thought patterns becoming “grooved” over time) or extrinsic causal processes, where genetic risk is mediated via the environment. An example of such a process would be the putatively causal link between cannabis use and schizophrenia (38,39). Cannabis is an environmental exposure, but its use is associated with genetic variants linked to impulsivity (40) and risk-taking (41). Assuming a demonstrable causal relationship between cannabis use and the development of schizophrenia, the environment (cannabis use) would *mediate* the effects of these genetic variants on schizophrenia; and impulsivity and risk-taking earlier in development would be on the causal pathway.

We applied structural models to associations between schizophrenia PRS and symptoms of childhood psychopathology to provide context that might render some of these competing explanations more or less likely. Using developmental models, we found that associations between schizophrenia PRS and emotional and behavioural symptomatology were best explained via effects on developmentally stable growth processes. This is in line with evidence on the stability of genetic influences, in general, across childhood (21). We had limited power to investigate the nature of these stable associations, but found evidence that, for behavioural problems, schizophrenia-associated alleles are primarily predictive of individual differences in their propensity to change over time in early childhood. Such a pattern may be more consistent with an aetiological model implicating environmentally mediated genetic effects and active or evocative gene-environment correlations, than direct pleiotropy. Future work incorporating measured environmental mediators may be able to establish whether these mechanisms are involved in the childhood manifestation of schizophrenia genetic liability.

The *“p”* factor structure of childhood psychopathology has been largely supported by previous genetically-informative work indicating that genetic prediction of symptoms is primarily mediated via a domain-general factor (e.g., (23)). Schizophrenia genetic risk has been shown to associate with psychopathology via this route (14,16). In apparent contrast, we found that models with symptom-specific effects were preferred, despite the cost of estimating many more parameters. There is likely some overfitting involved in these models, which is why we do not interpret the individual symptom-level effects. Nonetheless, the heterogeneity of these effects – even within domains – is clear and strengthens the conclusion that genetic liability for schizophrenia may influence childhood psychopathology in a more specific and less generalized manner than has been shown previously.

It is likely that two aspects of our approach explain these differences. First, we used item-level data, providing more scope for heterogeneity – from both signal and noise – to emerge, whereas scale-level data has often been used in the past. Second, we tested schizophrenia PRS at different thresholds – including thresholds at which all or almost all SNPs are included in the score. Previous work has often selected a single threshold score, typically *p*< 0.05 or lower. We found that lower threshold scores were likely to have smaller associations that, partly for reasons of statistical power, led to more parsimonious models being preferred. However, the amount of variance explained in symptoms continued to increase for scores created with less stringent thresholds suggesting that a single-score approach to using PRS potentially leads to meaningful signal from SNPs being missed. Using scores at higher thresholds both increases power and is consistent with the theory underpinning the polygenic model of genetic effects (5).

The small effect sizes for schizophrenia PRS on measures of childhood psychopathology in our sample are consistent with the literature (9,12,15). Our latent profile analytic approach demonstrates one way in which combining information across time and symptom domains can draw out more meaningful effect sizes (i.e., we observed effects equivalent to a 30% increase in the probability of displaying a particular symptom profile per standard deviation increase in schizophrenia PRS). However, we acknowledge the inherent bounds on the utility of effects explaining so little variance in childhood and adolescent outcomes. Using PRS for adult psychiatric disorders to identify children and adolescents at substantially increased, personal risk for behavioural and emotional difficulties and basing targeted prevention strategies on this information is currently not a realistic prospect. If the true effects captured by PRS are of the magnitude that the evidence so far suggests, PRS will have more value as tools to help clarify the mechanisms by which symptoms emerge, are maintained, differentiate, exacerbate or improve developmentally, than as individual-level predictors of disorder risk.

Our study is subject to several limitations. Small effect sizes for schizophrenia PRS on childhood traits mean that, even with a sample of several thousand children, we have limited power for differentiating similar parameterisations (e.g., on latent growth processes) and therefore opted against stratifying by sex. Secondly, the genotyped sample we use is subject to some selection effects, and the MoBa sample as a whole is (like all cohort studies) affected by selective attrition as the propensity to drop out of studies over time is linked to poorer health, which is also reflected genetically (42). Furthermore, the measures used to index symptoms of emotional and behavioural psychopathology are relatively brief and available only as maternal reports. Corroboration by additional reporters and using clinical interviews or diagnoses from linked health care registries would help to mitigate this limitation in future work.

Overall, our findings suggest that schizophrenia-related genetic variants are associated with symptoms of psychopathology stably from as early as 18 months of age, and with considerable phenotypic specificity by middle childhood. Further work – such as expanding models longitudinally and to incorporate measured environments and testing within-family polygenic prediction – can help to refine and triangulate developmental explanations for the emergence of schizophrenia later in life.

## Data Availability

The consent given by the participants does not open for storage of data on an individual level in repositories or journals. Researchers who want access to data sets for replication should submit an application to. Access to data sets requires approval from The Regional Committee for Medical and Health Research Ethics in Norway and an agreement with MoBa.

## Author contributions

LJH and AH conceived of the analyses; LJH, RBA, HA, and AH refined the analysis plan, prepared the polygenic scores and conducted the analyses; LJH, EC, and ØH performed quality control of the genetic data; PMM, PN, OAA, and TRK carried out the genotyping; LJH and AH drafted the manuscript; all authors critically reviewed and commented upon the manuscript.

## Acknowledgements

We are grateful to all the participating families in Norway who take part in this on-going cohort study.

The Norwegian Mother, Father, and Child Cohort Study is supported by the Norwegian Ministry of Health and Care Services and the Ministry of Education and Research.

We thank the Norwegian Institute of Public Health (NIPH) for generating high-quality genomic data. This research is part of the HARVEST collaboration, supported by the Research Council of Norway (NRC) (#229624). We also thank the NORMENT Centre for providing genotype data, funded by NRC (#223273), South East Norway Health Authority and KG Jebsen Stiftelsen. Further we thank the Center for Diabetes Research, the University of Bergen for providing genotype data and performing quality control and imputation of the data.

## References

1. Gratten J, Wray NR, Keller MC, Visscher PM. Large-scale genomics unveils the genetic architecture of psychiatric disorders. Nat Neurosci. 2014;17(6):782–90.

2. Schizophrenia Working Group of the Psychiatric Genomics Consortium, Ripke S, Neale BM, Corvin A, Walters JTR, Farh K-H, et al. Biological insights from 108 schizophrenia-associated genetic loci. Nature. 2014 Jul 22;511(7510):421–7.

3. Hirschhorn JN, Daly MJ. Genome-wide association studies for common diseases and complex traits. Nat Rev Genet. 2005;6(2):95–108.

4. Lam M, Chen CY, Li Z, Martin AR, Bryois J, Ma X, et al. Comparative genetic architectures of schizophrenia in East Asian and European populations. Nat Genet. 2019;51(12):1670–8.

5. Dudbridge F. Power and Predictive Accuracy of Polygenic Risk Scores. PLoS Genet. 2013;

6. Mistry S, Harrison JR, Smith DJ, Escott-Price V, Zammit S. The use of polygenic risk scores to identify phenotypes associated with genetic risk of schizophrenia: Systematic review. Schizophr Res. 2018 Jul;197(November 2017):2–8.

7. Kessler RC, Angermeyer M, Anthony JC, de Graaf R, Demyttenaere K, Gasquet I, et al. Lifetime prevalence and age-of-onset distributions of mental disorders in the World Health Organization’s. World Psychiatry 2007;6:168–176). 2007;6(October):168-76.

8. Serdarevic F, Jansen PR, Ghassabian A, White T, Jaddoe VW V., Posthuma D, et al. Association of Genetic Risk for Schizophrenia and Bipolar Disorder With Infant Neuromotor Development. JAMA Psychiatry. 2018 Jan 1;75(1):96.

9. Riglin L, Collishaw S, Richards A, Thapar AK, Maughan B, O’Donovan MC, et al. Schizophrenia risk alleles and neurodevelopmental outcomes in childhood: a population-based cohort study. The Lancet Psychiatry. 2017;4(1):57–62.

10. Reed ZE, Jones HJ, Hemani G, Zammit S, Davis OSP. Schizophrenia liability shares common molecular genetic risk factors with sleep duration and nightmares in childhood {version 2; peer review: 2 approved}. Wellcome Open Res. 2019;4:1–25.

11. Germine L, Robinson EB, Smoller JW, Calkins ME, Moore TM, Hakonarson H, et al. Association between polygenic risk for schizophrenia, neurocognition and social cognition across development. Transl Psychiatry. 2016;6(10):e924.

12. Nivard MG, Gage SH, Hottenga JJ, Van Beijsterveldt CEM, Abdellaoui A, Bartels M, et al. Genetic Overlap between Schizophrenia and Developmental Psychopathology: Longitudinal and Multivariate Polygenic Risk Prediction of Common Psychiatric Traits during Development. Schizophr Bull. 2017;43(6):1197–207.

13. Riglin L, Collishaw S, Richards A,… AKT-P, undefined 2017. The impact of schizophrenia and mood disorder risk alleles on emotional problems: investigating change from childhood to middle age. Psychol Med. 2017;

14. Riglin L, Thapar AK, Leppert B, Martin J, Richards A, Anney R, et al. The contribution of psychiatric risk alleles to a general liability to psychopathology in early life. bioRxiv. 2018;409540.

15. Jansen PR, Polderman TJC, Bolhuis K, van der Ende J, Jaddoe VWV, Verhulst FC, et al. Polygenic scores for schizophrenia and educational attainment are associated with behavioural problems in early childhood in the general population. J Child Psychol Psychiatry Allied Discip. 2018;59(1):39–47.

16. Jones HJ, Heron J, Hammerton G, Stochl J, Jones PB, Cannon M, et al. Investigating the genetic architecture of general and specific psychopathology in adolescence. Transl Psychiatry. 2018 Dec 8;8(1):145.

17. Jones HJ, Stergiakouli E, Tansey KE, Hubbard L, Heron J, Cannon M, et al. Phenotypic manifestation of genetic risk for schizophrenia during adolescence in the general population. JAMA Psychiatry. 2016;73(3):221–8.

18. Solmi F, Mascarell MC, Zammit S, Kirkbride JB, Lewis G. Polygenic risk for schizophrenia, disordered eating behaviours and body mass index in adolescents. Br J Psychiatry. 2019;215(1):428–33.

19. Hiemstra M, Nelemans SA, Branje S, van Eijk KR, Hottenga JJ, Vinkers CH, et al. Genetic vulnerability to schizophrenia is associated with cannabis use patterns during adolescence. Drug Alcohol Depend. 2018;

20. Magnus P, Birke C, Vejrup K, Haugan A, Alsaker E, Daltveit AK, et al. Cohort Profile Update: The Norwegian Mother and Child Cohort Study (MoBa). Int J Epidemiol. 2016 Apr;45(2):382–8.

21. Hannigan LJ, Walaker N, Waszczuk MA, McAdams TA, Eley TC. Aetiological Influences on Stability and Change in Emotional and Behavioural Problems across Development: A Systematic Review. Psychopathol Rev. 2017 Mar 21;a4(1):52–108.

22. Caspi A, Houts RM, Belsky DW, Goldman-Mellor SJ, Harrington H, Israel S, et al. The p Factor. Clin Psychol Sci. 2014 Mar 14;2(2):119–37.

23. Selzam S, Coleman JRI, Caspi A, Moffitt TE, Plomin R. A polygenic p factor for major psychiatric disorders. Transl Psychiatry. 2018;8(1):205.

24. Allegrini AG, Cheesman R, Rimfeld K, Selzam S, Pingault J-B, Eley T, et al. The p factor: Genetic analyses support a general dimension of psychopathology in childhood and adolescence. bioRxiv. 2019;591354.

25. Riglin L, Thapar AK, Leppert B, Martin J, Richards A, Anney R, et al. Using Genetics to Examine a General Liability to Childhood Psychopathology. Behav Genet. 2019;(0123456789).

26. Leppert B, Havdahl A, Riglin L, Jones HJ, Zheng J, Davey Smith G, et al. Association of maternal neurodevelopmental risk alleles with early-life exposures. JAMA Psychiatry. 2019;76(8):834–42.

27. Magnus P, Irgens LM, Haug K, Nystad W, Skjærven R, Stoltenberg C, et al. Cohort profile: The Norwegian Mother and Child Cohort Study (MoBa). Int J Epidemiol. 2006;35(5):1146–50.

28. Achenbach TM, Ruffle TM. The Child Behavior Checklist and Related Forms for Assessing Behavioral/Emotional Problems and Competencies. Pediatr Rev. 2007;21(8):265–71.

29. Angold A, Costello EJ, Messer SC, Pickles A, Winder F, Silver D. The development of a short questionnaire for use in epidemiological studies of depression in children and adolescents. Int J Methods Psychiatr Res. 1995;5:237–49.

30. Birmaher B, Khetarpal S, Brent D, Cully M, Balach L, Kaufman J, et al. The Screen for Child Anxiety Related Emotional Disorders (SCARED): Scale construction and psychometric characteristics. J Am Acad Child Adolesc Psychiatry. 1997;36(4):545–53.

31. Silva RR, Alpert M, Pouget E, Silva V, Trosper S, Reyes K, et al. A rating scale for disruptive behavior disorders, based on the DSM-IV item pool. Psychiatr Q. 2005;76(4 SPEC. ISS.):327–39.

32. Euesden J, Lewis CM, O’Reilly PF. PRSice: Polygenic Risk Score software. Bioinformatics. 2015;

33. Lo Y, Mendell NR, Rubin DB. Testing the number of components in a normal mixture. Biometrika. 2001;

34. Vuong QH. Likelihood Ratio Tests for Model Selection and Non-Nested Hypotheses. Econometrica. 1989;

35. Rosseel Y. lavaan: An R Package for Structural Equation Modeling. J Stat Softw. 2015;

36. Muthén LK, Muthén BO. Mplus User’s Guide. Eighth Edi. 1998–2017. Los Angeles, CA: Muthén & Muthén;

37. Hallquist MN, Wiley JF. MplusAutomation: An R Package for Facilitating Large-Scale Latent Variable Analyses in Mplus. Struct Equ Model. 2018;

38. Vaucher J, Keating BJ, Lasserre AM, Gan W, Lyall DM, Ward J, et al. Cannabis use and risk of schizophrenia: A Mendelian randomization study. Mol Psychiatry. 2018;23(5):1287–92.

39. Gage SH, Jones HJ, Burgess S, Bowden J, Davey Smith G, Zammit S, et al. Assessing causality in associations between cannabis use and schizophrenia risk: A two-sample Mendelian randomization study. Psychol Med. 2017;47(5):971–80.

40. Soler Artigas M, Sánchez-Mora C, Rovira P, Richarte V, Garcia-Martínez I, Pagerols M, et al. Attention-deficit/hyperactivity disorder and lifetime cannabis use: genetic overlap and causality. Mol Psychiatry. 2019;

41. Strawbridge RJ, Ward J, Cullen B, Tunbridge EM, Hartz S, Bierut L, et al. Genome-wide analysis of self-reported risk-taking behaviour and cross-disorder genetic correlations in the UK Biobank cohort. Transl Psychiatry. 2018;8(1):1–11.

42. Adams MJ, Hill WD, Howard DM, Dashti HS, Davis KAS, Campbell A, et al. Factors associated with sharing e-mail information and mental health survey participation in large population cohorts. Int J Epidemiol. 2019 Jul 1;

